# Quantifying the Pharyngeal Wall Thickness on Modified Barium Swallow Studies: A Scoping Review and Reliability Study

**DOI:** 10.64898/2026.10.17.26365811

**Authors:** Shitong Mao, Holly McMillan, Jake D. Leone, Carly E.A. Barbon, Sheila Nida Buoy, Kristy Brock, Stephen Lai, Mohamed A Naser, Clifton David Fuller, Katherine A. Hutcheson, InHANCE Research Program Working Group

## Abstract

**Purpose:** Posterior pharyngeal wall (PPW) thickening is a common structural manifestation of treatment-related internal lymphedema in patients with head and neck cancer and can be evaluated using lateral videofluoroscopic images acquired during modified barium swallow (MBS) studies. However, no standardized method currently exists for quantitative measurement.

**Methods:** This two-phase methodological study first conducted a scoping review to characterize existing approaches to PPW thickness measurement and identify the principal factors contributing to variation in PPW thickness measurement. Eight studies were identified with variation observed across these measurement factors. An empirical reliability study was then performed using 107 MBS studies from 93 Head and Neck Cancer patients. Two independent raters annotated 10 anatomical landmarks on pre-swallow and post-swallow resting frames, and a computer-assisted algorithm quantitatively generated seven candidate PPW thickness measurements based on different vertebral reference points and measurement directions. Reliability was assessed intraclass correlation coefficients (ICCs).

**Results:** Inter-rater reliability was highest for the measurement originating from the midpoint of C3 and extending perpendicular to the anterior C3 surface (ICC, 0.78; 95% CI, 0.72-0.83). Among the normalization references, C2-C4 vertebral length demonstrated greater reliability than C3 vertebral length, with an ICC of 0.88 (95% CI, 0.85-0.91). No significant differences were observed between pre-swallow and post-swallow PPW thickness measurements across the evaluated approaches.

**Conclusions:** These findings provide a methodological foundation for more standardized and reliable quantification of PPW thickness, with potential to expand the role of MBS from the functional evaluation of swallowing to include quantitative characterization of pharyngeal structural abnormalities.

## Introduction

Advances in multimodality treatment have improved disease control and survival in patients with head and neck cancer (HNC), while increasing the burden of chronic treatment-related morbidity(1–4). Among these late effects, head and neck lymphedema (HNL) is common. HNL involves both external tissues of the face and neck, as well as the internal structures of the oral cavity, pharynx, and/or larynx (5–7). Internal lymphedema may develop or persist after treatment and contribute to long-term morbidity(5, 8). It has been associated with swallowing impairment, including penetration-aspiration, pharyngeal residue, diet modification, and patient-reported dysphagia(9–11). The posterior pharyngeal wall (PPW) is a key component of pharyngeal swallowing physiology, contributing to pharyngeal constriction and bolus clearance(12, 13). Treatment-induced lymphedema and fibrosis involving the PPW threatens to compromise these functions and contribute to chronic swallowing impairment(14, 15). Quantitative assessment of PPW thickness is a relatively simple objective structural measure reported in the literature as a method to quantify internal lymphedema in this population, feasible to collect on videofluoroscopy which is commonly performed to assess swallowing dysfunction in HNC survivors.

Imaging assessment of internal lymphedema has primarily relied on endoscopy and cross-sectional imaging modalities, including CT and MRI(6, 11, 16). Flexible transnasal endoscopic examination, including flexible nasendoscopy, fiberoptic laryngoscopy, and FEES (fiberoptic endoscopic evaluation of swallowing), is widely used to grade internal lymphedema across multiple pharyngolaryngeal sites, including the base of tongue, epiglottis, arytenoids, laryngeal folds, posterior pharyngeal wall, valleculae, and pyriform sinuses(7, 10, 17, 18). However, although the luminal surface of the PPW can be visually graded, endoscopy is incapable of delineate its full tissue depth or its spatial distance to the cervical spine and therefore does not permit direct quantification of PPW thickness. CT and MRI enable detailed cross-sectional assessment of pharyngeal soft tissues, whereas modified barium swallow (MBS) study offers a distinct opportunity to evaluate the PPW within the clinical workflow of swallowing assessment(19). During routine interpretation of MBS, speech-language pathologists assess swallowing physiology while simultaneously visualizing the PPW and adjacent cervical vertebral structures on lateral videofluoroscopic images. This concurrent visualization allows structural characteristics of the PPW to be examined without separating them from the swallowing events and functional measures to which they are clinically relevant. Quantitative analysis of PPW thickness on MBS therefore provides a framework for integrating structural assessment with established measures of swallowing safety, efficiency, and bolus clearance within the unique examination.

Some studies have recognized the potential of videofluoroscopic images acquired during MBS for structural analysis of the pharynx and adjacent cervical anatomy, and have applied these images to assess internal tissue changes, including PPW(20, 21). However, videofluoroscopic imaging presents inherent constraints when used for quantitative structural evaluation. MBS is a dynamic, real-time X ray examinations of swallowing examination in which image acquisition must balance adequate visualization with minimization of radiation exposure within ALARA standards (as low as reasonably achievable), particularly in patients with HNC who have frequently undergone prior radiotherapy and experienced treatment-related functional impairment(22). As a result, image quality and soft-tissue contrast may be limited, and the intensity transitions that define anatomical boundaries can be subtle, increasing uncertainty in visual delineation of the PPW(23, 24). PPW thickness measurements therefore require carefully selected anatomical landmarks and predefined measurement directions to reduce observer-dependent variation. In addition, unlike CT, videofluoroscopic imaging does not provide a fixed spatial scale because image magnification varies with the relative positions of the patient, X-ray source, and image receptor. Absolute distance measurements in millimeters or centimeters are therefore not directly available unless an external radiopaque calibration object, such as a coin of known dimensions, is included in the field of view. When such calibration is unavailable, anatomical reference lengths must be used for normalization. Existing studies have adopted different approaches to PPW thickness measurement on videofluoroscopic images from MBS, but these approaches have not been systematically compared, and their methodological suitability remains insufficiently examined.

To address these methodological uncertainties, we conducted a two-phase investigation comprising a scoping review and a quantitative reliability study. The scoping review aimed to identify the general approaches used to measure PPW thickness on MBS or other videofluoroscopic images and to determine the methodological variables that could affect the resulting measurements. Guided by this synthesis, we developed a computer-assisted, semi-automated approach using MBS examinations and assessed the comparative reliability of alternative anatomical reference points, measurement orientations, normalization scalars, and pre-versus post-swallow resting frames. Collectively, these analyses were intended to establish a reproducible and optimal annotation framework for quantitative PPW thickness assessment on MBS.

## Method

### 2.1 Scope review

As a formative step in the development of quantitative analysis, we conducted a structured literature review to identify previously reported approaches to PPW thickness measurement on lateral fluoroscopic images and to derive candidate methods for subsequent comparison. The literature search was conducted in four electronic databases—PubMed/MEDLINE, Web of Science Core Collection, Scopus, and Google Scholar. The search strategy combined keywords/ terminologies related to fluoroscopic swallowing imaging, including “videofluoroscopic swallowing study,” “videofluoroscopy,” “modified barium swallow,” “fluoroscopic swallowing study,” and “VFSS,” with keywords related to the anatomical structure and measurement of interest, including “posterior pharyngeal wall,” “pharyngeal wall thickness,” “pharyngeal thickness,” “pharyngeal soft tissue thickness,” and “PPW.”

Inclusion criteria were studies published after 2010 that involved human participants, used lateral fluoroscopic imaging, and reported quantitative measurement of PPW or prevertebral soft-tissue thickness. No restrictions were placed on clinical population or fluoroscopic task; eligible studies therefore included swallowing studies, non-swallowing motion studies, cervical spine imaging, assessments related to anterior cervical spine surgery, and studies of non-oncologic populations. Exclusion criteria were studies that did not use fluoroscopic imaging, did not quantify PPW thickness, used only non-lateral projections, provided insufficient methodological detail, combined fluoroscopic measurements with other imaging modalities without separately interpretable results, or were reviews, conference abstracts, case reports, pediatric studies, or studies based exclusively on animal or cadaveric data.

For each study, the PPW thickness measurement approach was evaluated with respect to the methodological components that could influence the resulting measurement: (1) bolus selection, characterized by volume and consistency; (2) frame selection, characterized by the criteria used to select the frame and whether a pre-swallow or post-swallow frame was used; (3) anatomical starting point and (4) measurement orientation, which defined the single measurement. MBS studies were acquired as routine, clinical procedures according to a standard protocol. The standard bolus protocol included 2 trials each of 5-mL, 10-mL, and self-administered sips from a cup of thin liquid barium (Varibar; Bracco Diagnostics, Inc), barium pudding (Varibar; Bracco Diagnostics, Inc), and a cracker coated in barium pudding.

### 2.2 Data collection and annotation

This cross-sectional methodological study involved a secondary analysis of de-identified MBS data collected from patients with HNC undergoing a radiation-based treatment pathway at University of Texas MD Anderson Cancer Center under an Institutional Review Board-approved protocol (PA11-0849), with the analysis focused on the evaluation of PPW thickness measurement methods. Eligible participants were adults (≥18 years) with a diagnosis of head and neck malignancy who were receiving curative-intent external beam radiotherapy of at least 60 Gy.

The MBS examinations were performed as part of routine clinical care using a standardized protocol. The bolus protocol included two trials for each of the following bolus conditions: 5-mL thin liquid barium, 10-mL thin liquid barium, self-administered cup sips of thin liquid barium, barium pudding (Varibar; Bracco Diagnostics, Inc.), and a cracker coated with barium pudding. MBS videos were recorded using the TIMS system (TIMS Medical, LLC, Chelmsford, MA, USA) at a frame rate of 30 frames per second (FPS). Only examinations with complete video acquisition were included. The recordings were stored in DICOM format, and only DICOM files that were complete and free of file corruption were retained for analysis. Examinations were further excluded if any object or imaging artifact obscured the cervical vertebrae or pharyngeal region required for PPW measurement. The study sample comprised 107 MBS examinations from 93 patients.

The MBS examinations stored in DICOM format were further converted to AVI video files while preserving the original frame rate and spatial resolution. Two independent raters reviewed all MBS examinations and manually measured PPW thickness using an open-source image analysis software ImageJ. All MBS videos included in the analysis were acquired in the lateral view. Although routine clinical MBS protocols also include bolus trials in the anterior-posterior view, these trials were excluded from this study. A 5-mL thin liquid bolus, the smallest liquid volume included in the standardized MBS protocol, was selected for PPW thickness measurement. As two 5-mL thin liquid trials were acquired for each examination, both were reviewed by the raters, and the trial providing superior visualization of the anatomical structures relevant to PPW measurement was selected for analysis. For the selected bolus trial, two frames were selected: one from the pre-swallow resting period and one from the post-swallow resting period. Within each period, the selected frame was the first frame in which the pyriform sinuses reached their lowest position relative to the cervical spine(25, 26).

### 2.3 A computer-assisted semi-automatic approach

A computer-assisted, semi-automated approach was developed to measure PPW thickness on the selected frames while minimizing the extent of manual annotation. For each frame, the rater manually identified five anterior cervical vertebral corners spanning C2-C4 and five corresponding points along the anterior boundary of the PPW. The PPW boundary points were placed to provide slightly greater superior-to-inferior coverage than the span defined by the vertebral corners, ensuring that the region required for subsequent measurements was encompassed. Thus, the manual annotation consisted of pinpointing 10 key-points in total (P1-P10), after which the computer-assisted procedure used these pixel-based coordinates to derive the PPW measurements, as shown in Figure 1. Detailed definitions of the 10 annotated landmarks are provided in Table 1.

**Figure 1.**
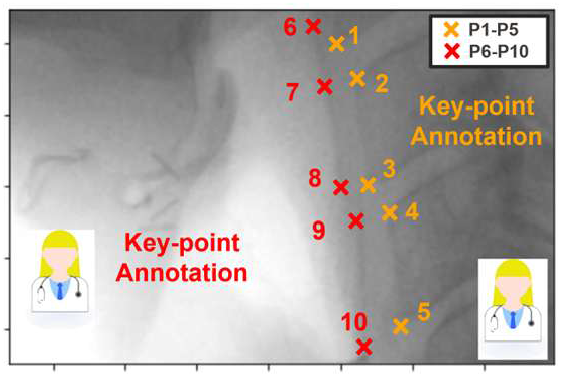
Manual annotation of five anterior cervical vertebral corners (P1-P5) and five corresponding PPW boundary points (P6-P10) for semi-automated PPW thickness measurement.

**Table 1.** Definitions of the 10 manually annotated key points used for semi-automated PPW thickness measurement.

| Point number | Definitions | Point number | Definitions |
| --- | --- | --- | --- |
| 1 | Anterior Inferior C2 corner | 6 | Anterior Boundary of PPW slightly superior to Point 1 |
| 2 | Anterior Superior C3 Corner | 7 | Anterior Boundary of PPW across from Point 2 |
| 3 | Anterior Inferior C3 Corner | 8 | Anterior Boundary of PPW across from Point 3 |
| 4 | Anterior Superior C4 Corner | 9 | Anterior Boundary of PPW across from Point 4 |
| 5 | Anterior Inferior C4 Corner | 10 | Anterior Boundary of PPW slightly inferior to Point 5 |

The computer-assisted measurement procedure is demonstrated in Figure 2 using geometric calculation. First, the coordinates of the five manually annotated PPW boundary points (P6-P10) were used to reconstruct the PPW contour. A cubic spline interpolation was applied to these points to generate a smooth curve passing through all five landmarks, as shown in Figure 2(a). The resulting curve was then densely sampled at 2,000 interpolated points, providing a continuous numerical representation of the

**Figure 2.**
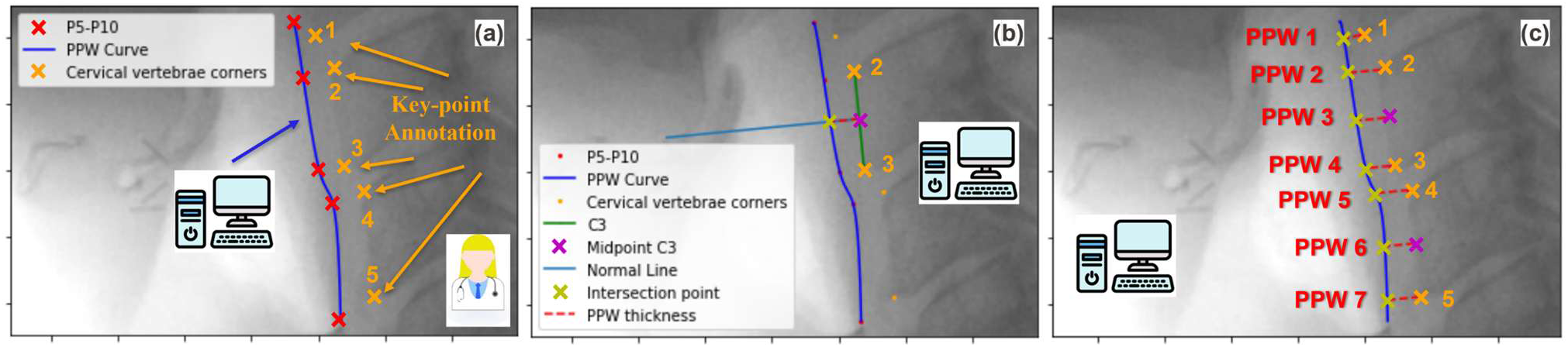
Semi-automated workflow for PPW thickness measurement. (a) PPW contour fitting based on P6-P10; (b) Example PPW measurement using the C3 midpoint as the starting point; (c) Seven candidate PPW measurement approaches.

#### PPW contour for subsequent distance measurements

As an example, Figure 2(b) illustrates the measurement procedure using the midpoint of the anterior C3 surface. The midpoint was calculated from the coordinates of the anterior-superior and anterior-inferior C3 corners (P2 and P3). From this midpoint, a normal line was extended toward the PPW contour in a direction perpendicular to the anterior C3 surface, represented by the line connecting P2 and P3. The intersection of this normal line with the reconstructed PPW contour was then identified, and PPW thickness was defined as the distance from the C3 midpoint to this intersection point. All calculations were performed computationally and automatically after completion of the manual key-point annotations.

Using similar computational procedures, seven candidate PPW thickness measurement approaches were conducted by different anatomical starting points and measurement directions, as shown in Figure 2(c). The specific definitions of these seven approaches are provided in Table 2. Additionally, the C2-C4 distance (P1-P5) and C3 distance (P2-P3) were also calculated as anatomical scalar lengths for normalization of PPW thickness measurements(27–29). All computational procedures were implemented in Python (version 3.10.10).

**Table 2.** Definitions of the seven candidate PPW thickness measurement approaches based on anatomical starting point and measurement direction.

| Options | Starting point | Direction- Perpendicular to the line connecting |
| --- | --- | --- |
| PPW 1 | Anterior <u>Inferior</u> C2 corner | C2C4 corners (P1-P5) |
| PPW 2 | Anterior <u>Superior</u> C3 Corner | C3 corners (P2-P3) |
| PPW 3 | <u>Mid-point</u> of C3 | C3 corners (P2-P3) |
| PPW 4 | Anterior <u>Inferior</u> C3 Corner | C3 corners (P2-P3) |
| PPW 5 | Anterior <u>Superior</u> C4 Corner | C4 corners (P4-P5) |
| PPW 6 | <u>Mid-point</u> of C4 | C4 corners (P4-P5) |
| PPW 7 | Anterior <u>Inferior</u> C4 Corner | C4 corners (P4-P5) |

**Table 3.**
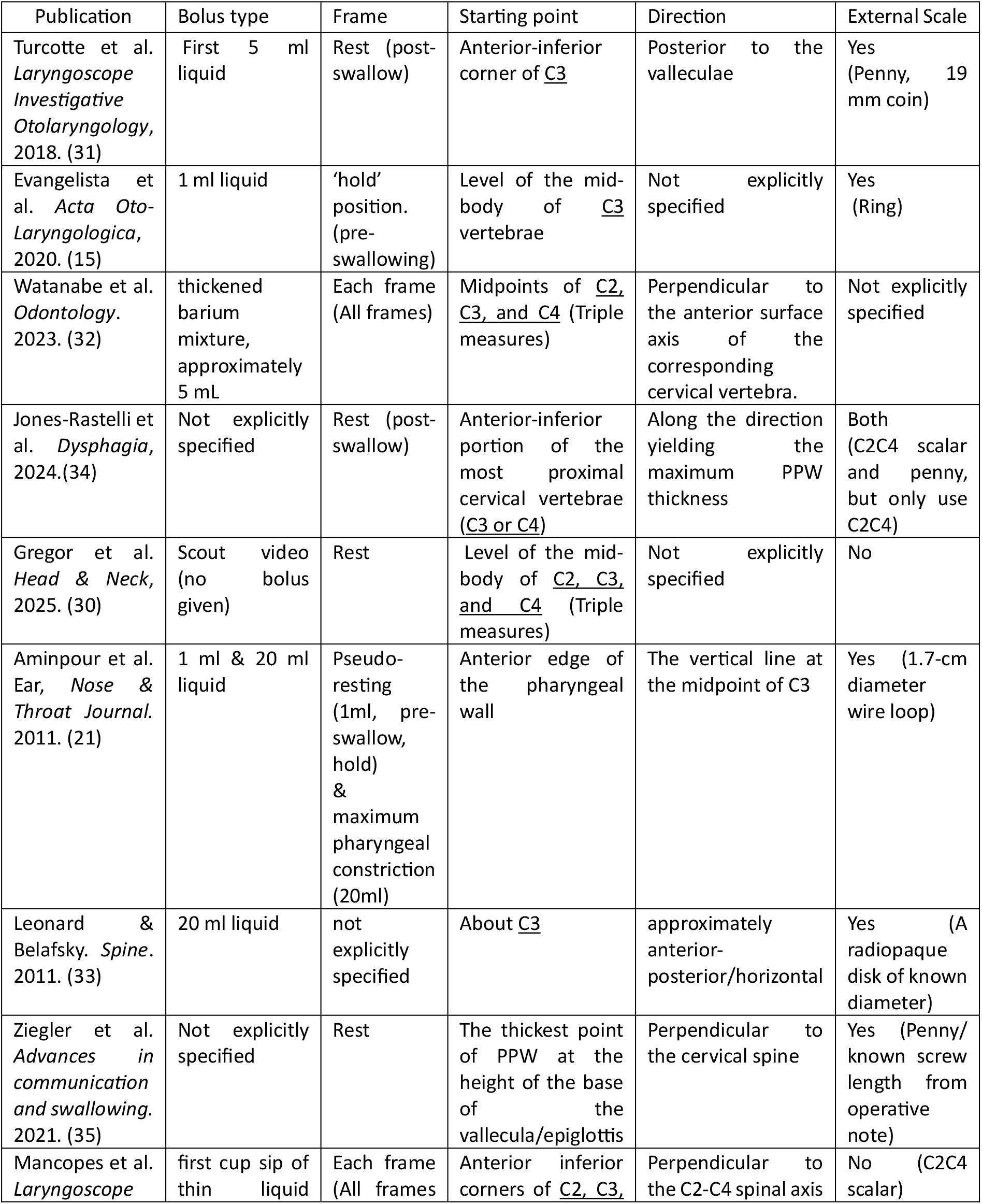
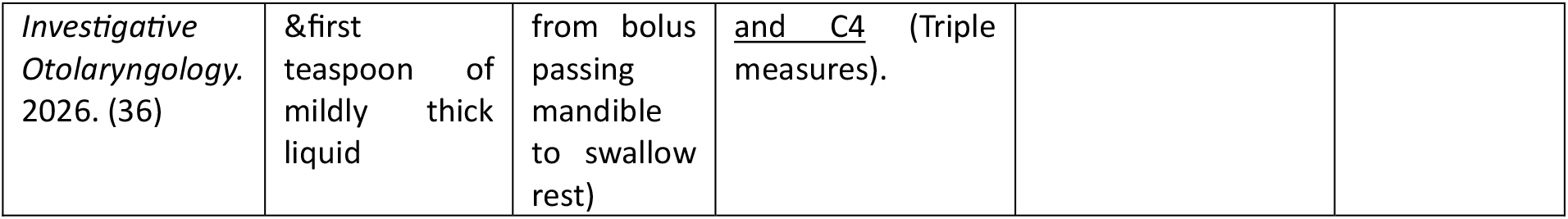
Methodological characteristics of PPW thickness measurement approaches across current studies.

### 2.4 Statistical Analysis

Statistical analyses were designed to evaluate the proposed PPW measurement framework: the interrater reliability of the different measurement approaches, the effect of anatomical normalization on measurement reliability, and the potential effect of frame selection on PPW thickness measurements. To assess measurement reliability, intraclass correlation coefficients (ICCs) were calculated between the two independent raters. ICCs were first determined for each of the seven PPW thickness measurement approaches derived from the raters’ independent key-point annotations. The reliability of the two anatomical scalar lengths, C2C4 and C3, was evaluated separately. Each of the seven PPW thickness measurements was then normalized using either the C2-C4 or C3 reference length, and ICCs were recalculated for the normalized measurements to examine the reliability of each normalization strategy.

The effect of frame selection on PPW thickness measurements was examined by comparing measurements obtained from the pre-swallow and post-swallow resting frames. Paired t-tests were performed for each of the seven measurement approaches. These comparisons were conducted separately for measurements from each rater and for the mean measurement across the two raters. Statistical significance was defined as p < 0.05. All statistical analyses were performed in Python (version 3.10.10) using SciPy (version 1.12.0).

## Results

### 3.1 General measurements of the PPW measurements

The scoping review identified nine representative studies that quantitatively assessed PPW thickness via MBS or other videofluoroscopic imaging. Across the included studies, PPW thickness was represented by a single straight-line measurement or a set of straight-line measurements on lateral videofluoroscopic images, rather than by segmentation of the entire PPW region. These measurements were not defined by specifying two fixed endpoints. Instead, all measurements were defined by an anatomical starting point, most commonly located on the cervical vertebrae, and a prescribed measurement direction. Along this direction, a line was then extended from the starting point toward the anterior PPW boundary, and its intersection with the visible soft-tissue boundary defined the second endpoint of the measurement. PPW thickness was quantified as the distance between these two points. The general concept was shown in Figure 3.

**Figure 3.**
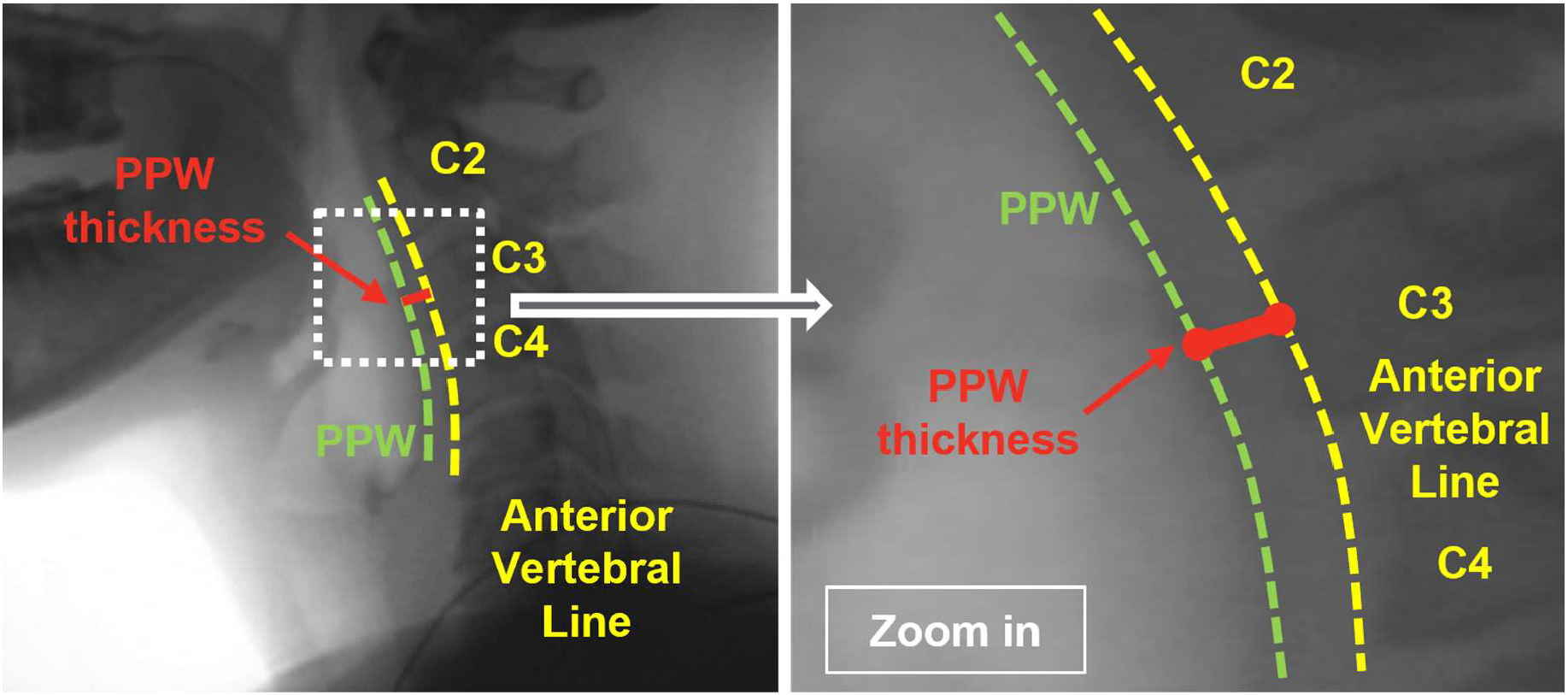
General concept of PPW thickness measurement on lateral videofluoroscopic imaging, defined as the distance from the anterior cervical vertebral reference line to the PPW boundary

### 3.2 Current PPW measurements

The included studies demonstrated variation in how PPW thickness was measured, as shown in **Error! Reference source not found.**. Although those studies were not limited to patients with HNC, all incorporated measurement of PPW thickness within their respective clinical or physiologic contexts. Gregor et al., Evangelista et al., and Turcotte et al. examined patients with HNC who had received radiation-based treatment, although cancer subsites and treatment pathways differed across studies(15, 30, 31). Gregor et al. was distinct in that no bolus was administered, and the videofluoroscopy was used solely as an imaging modality to evaluate immediate changes in internal edema following a single pneumatic compression treatment(30). Watanabe et al. studied postoperative tongue cancer patients treated with surgery alone, without chemotherapy or radiation therapy(32). Leonard & Belafsky, Jones-Rastelli et al., and Ziegler et al. studied patients undergoing anterior cervical spine surgery, with particular emphasis on postoperative dysphagia or surgery-related changes in swallowing(33–35). Aminpour et al. and Mancopes et al. evaluated PPW characteristics in healthy adults, with emphasis on age-related differences in PPW thickness and swallowing physiology(21, 36). In addition to static PPW thickness measurements,

Mancopes et al. and Watanabe et al. also performed dynamic analyses of PPW movement(32, 36).

Bolus selection varied across studies. Because MBS examinations typically comprise a series of bolus trials, PPW thickness was not measured from every trial; instead, studies selected a specific bolus trial for measurement, except in studies that used videofluoroscopy for imaging without bolus administration(30). Among the nine studies, five explicitly used a liquid bolus. Of the studies reporting bolus volume, two used 1 mL, two used 5 mL, and two used 20 ml. When more than one trial was available with the same bolus volume and consistency, the first trial was selected for measurement(31, 36).

Frame selection varied according to the purpose of PPW thickness measurement. Studies focused on pharyngeal wall movement generally perform measurements across all video frames. Two of the nine included studies used this frame-selection strategy(32, 36). Most other studies selected a relatively static frame for measurement, although one study evaluated PPW thickness at maximum pharyngeal constriction(21). Among studies using static frames, measurements were obtained at a resting state either before or after the swallow. Two studies explicitly used a pre-swallow “hold” position(15, 21), whereas two others specified a post-swallow resting frame(31, 34). The latter studies provided similar operational definitions of post-swallow rest, with the resting frame defined as the frame of lowest pyriform sinuses position after the epiglottis had returned to rest(25, 26).

Most studies used the cervical vertebrae as the anatomical reference for initiating PPW thickness measurement, while only two of the nine studies began from the PPW and extended the measurement toward the cervical vertebrae(21, 35). In one of the PPW-based approaches, Ziegler et al. identified the thickest portion of the PPW near the base of the vallecula/epiglottis. In one of the studies with the starting point located on the PPW, Ziegler JP et al. identified the thickest portion of the PPW near the base of the vallecula/epiglottis(35). For measurements initiated from the cervical vertebrae, the starting point was typically defined using an anterior bony landmark visible on lateral videofluoroscopy. Three studies conducted measurements at three cervical locations for triple measures, using C2, C3, and C4 as references(30, 32, 36). Watanabe et al. used the midpoints of these vertebral levels(32), whereas Mancopes et al. used the anterior-inferior corners(36). Other studies performed a single measurement at one cervical level. Three of these studies used C3 as the reference for starting points, with Turcotte et al. specifically defining the starting point as the anterior-inferior corner of C3(15, 31, 33). The study of Jones-Rastelli et al. used a distinct strategy in which the thickest portion of the PPW was first identified, followed by selection of the nearest cervical vertebral level, and the PPW thickness was then measured between the PPW and the anterior-inferior portion of that vertebra(34). Depending on the location of the thickest PPW segment, the corresponding vertebral reference could be at C3 or C4. This strategy was conceptually similar to that of Ziegler JP., as both focused on the thickest portion of the PPW(35).

Three studies explicitly specified that the measurement line extending from the starting point(s) toward the PPW should be perpendicular to a defined anatomical reference. Watanabe et al. conducted three measurements at C2, C3, and C4, with each measurement direction defined as perpendicular to the anterior surface of the corresponding vertebra, represented by the line connecting its anterior-superior and anterior-inferior corners separately(32). Mancopes et al. also measured at C2, C3, and C4, but all three measurement lines were oriented perpendicular to a common spinal axis, defined as the line extending from the anterior-inferior corner of C2 to the anterior-inferior corner of C4, and the three measurement lines were parallel to one another(36). Ziegler et al. defined the measurement direction as perpendicular to a line formed by the anterior surfaces of C2, C3 and C4, a reference that was likely facilitated by the postoperative cervical alignment in their anterior cervical spine surgery cohort(35).

Five studies explicitly used an external object of known size as a reference, allowing PPW thickness measurements to be scaled and reported as absolute distances (cm or mm). Three studies instead used the C2-C4 length as an internal anatomical scalar. Jones-Rastelli et al. included an external penny but normalized PPW thickness to C2-C4, and the penny was used only to validate the stability of the C2-C4 reference across timepoints, with no within-subject change observed in penny size when expressed relative to C2-C4(34). Gregor et al. likewise used C2-C4 as an internal reference, but they adjusted image size to generate identical C2-C4 length in pixel at each timepoint(30). Mancopes et al. explicitly used the C2-C4 distance as the normalization scalar(36).

### 3.3 Reliability analysis of computer-assisted PPW measurements

For each selected bolus trial, both the pre-swallow and post-swallow resting frames were independently annotated by each rater. This resulted in 214 annotation sets per rater across the 107 MBS examinations. Each annotation set generated PPW thickness measurements automatically for all seven candidate approaches using the computer-assisted methods, and interrater reliability was then evaluated by comparing the corresponding measurements derived from the two raters using ICCs. Interrater reliability was first assessed using the PPW thickness measurements without normalization, expressed directly in pixels. The corresponding ICC results are presented in Table 4.

**Table 4.** Interrater reliability of the seven PPW thickness measurement approaches and cervical vertebral reference lengths.

| Approach | ICC (95% CI) (PPW thickness in pixels) |
| --- | --- |
| PPW 1 | 0.704 (0.630, 0.766) |
| PPW 2 | 0.733 (0.665, 0.790) |
| PPW 3 | **0.783 (0.725, 0.830)** |
| PPW 4 | **0.756 (0.692, 0.808)** |
| PPW 5 | 0.623 (0.534, 0.699) |
| PPW 6 | 0.621 (0.531, 0.697) |
| PPW 7 | 0.508 (0.402, 0.601) |
| C2-C4 length | **0.883 (0.850, 0.910)** |
| C3 length | 0.719 (0.647, 0.778) |

Measurements with starting points at C3 showed relatively higher interrater reliability. PPW 3, with the C3 midpoint as the starting point, demonstrated the highest interrater reliability among the seven measurement approaches (ICC = 0.78, 95% CI: 0.72-0.83), followed by PPW 4, with the anterior-inferior C3 corner as the starting point (ICC = 0.76, 95% CI: 0.69-0.81), and PPW 2, with the anterior-superior C3 corner as the starting point (ICC = 0.73, 95% CI: 0.66-0.79). PPW 5-7, with starting points at C4, showed lower interrater reliability, with ICCs ranging from 0.51 to 0.62. The C2-C4 reference length showed a higher ICC of 0.88 (95% CI: 0.85-0.91), compared with 0.72 (95% CI: 0.65-0.78) for the C3 reference length.

Pairwise comparisons of the seven pixel-based PPW thickness measurement approaches are shown in Table 5. Significant differences in interrater reliability were primarily observed between the higher-reliability C3-based approaches (PPW2-4) and the lower-reliability C4-based approaches (PPW5-7), whereas differences among the C2- and C3-based approaches were generally not significant.

**Table 5.** Pairwise ICC differences among the seven pixel-based PPW thickness measurement approaches.

| Approach | PPW 2 | PPW 3 | PPW 4 | PPW 5 | PPW 6 | PPW 7 |
| --- | --- | --- | --- | --- | --- | --- |
| PPW 1 | $\Delta = -0.029$<br>[-0.109, 0.047] | **$\Delta = -0.078$**<br>**[-0.162, -0.004]** | $\Delta = -0.052$<br>[-0.150, 0.033] | $\Delta = -0.081$<br>[-0.017, 0.182] | $\Delta = -0.084$<br>[-0.013, 0.198] | **$\Delta = 0.196$**<br>**[0.063, 0.304]** |
| PPW 2 | | $\Delta = -0.049$<br>[-0.111, 0.003] | $\Delta = -0.023$<br>[-0.107, 0.063] | **$\Delta = 0.110$**<br>**[0.036, 0.199]** | **$\Delta = 0.112$**<br>**[0.021, 0.226]** | **$\Delta = 0.225$**<br>**[0.090, 0.388]** |
| PPW 3 | | | $\Delta = 0.026$<br>[-0.012, 0.073] | **$\Delta = 0.159$**<br>**[0.090, 0.241]** | **$\Delta = 0.162$**<br>**[0.084, 0.265]** | **$\Delta = 0.274$**<br>**[0.142, 0.424]** |
| PPW 4 |  |  |  | **$\Delta = 0.133$**<br>**[0.053, 0.222]** | **$\Delta = 0.135$**<br>**[0.056, 0.239]** | **$\Delta = 0.248$**<br>**[0.125, 0.377]** |
| PPW 5 | | | | | $\Delta = 0.003$<br>[-0.062, 0.070] | **$\Delta = 0.115$**<br>**[0.003, 0.235]** |
| PPW 6 |  |  |  |  |  | **$\Delta = 0.113$**<br>**[0.043, 0.185]** |
| $\Delta = ICC_{row} - ICC_{column}$, presenting the ICC difference. A positive $\Delta$ indicates higher reliability for the row measurement, and vice versa for a negative $\Delta$. A 95% CI including 0 indicates no significant difference; conversely, a CI excluding 0 indicates a significant difference (highlighted cells). | | | | | | |

Normalization using the C2-C4 length generally yielded higher interrater reliability than normalization using the C3 reference length, as shown in Table 6. With C2-C4 normalization, PPW 3 (starting at the C3 midpoint) showed the highest ICC (0.77, 95% CI: 0.70-0.82). PPW 1, PPW 2, and PPW 4 also showed ICCs above 0.70, ranging from 0.73 to 0.75. With C3 length normalization, PPW 3 remained the highest (ICC = 0.71, 95% CI: 0.63-0.77), followed by PPW 2 (starting point at the C3 superior corner, ICC = 0.70, 95% CI: 0.63-0.76). Consistent with the results without the normalization, PPW 5-7 showed lower ICCs under both normalization strategies.

**Table 6.** Interrater reliability of PPW thickness measurements normalized by C2-C4 and C3 reference lengths.

| Approach | ICC (95% CI), PPW thickness normalized by |  | ICC comparison |  |
| --- | --- | --- | --- | --- |
| | C3 length | C2C4 length | ICC difference<br>$\Delta = ICC_{C3} - ICC_{C2C4}$ | 95% CI of $\Delta ICC$ |
| PPW 1 | 0.685 (0.607, 0.750) | 0.753 (0.688, 0.805) | **-0.067** | **[-0.127, -0.013]** |
| PPW 2 | 0.702 (0.627, 0.764) | 0.734 (0.67, 0.79) | -0.033 | [-0.084, 0.014] |
| PPW 3 | **0.707 (0.633, 0.768)** | **0.766 (0.704, 0.816)** | **-0.059** | **[-0.059, -0.113]** |
| PPW 4 | 0.663 (0.581, 0.732) | 0.737 (0.669, 0.792) | **-0.074** | **[-0.074, -0.131]** |
| PPW 5 | 0.561 (0.462, 0.647) | 0.603 (0.511, 0.682) | **-0.042** | **[-0.042, -0.088]** |
| PPW 6 | 0.621 (0.532, 0.697) | 0.647 (0.562, 0.719) | **-0.026** | **[-0.026, -0.075]** |
| PPW 7 | 0.576 (0.479, 0.659) | 0.563 (0.464, 0.648) | 0.014 | [0.014, -0.043] |

**Table 7.** Pairwise ICC differences among the seven PPW thickness measurement approaches normalized by C3 length.

| Approach | PPW 2 | PPW 3 | PPW 4 | PPW 5 | PPW 6 | PPW 7 |
| --- | --- | --- | --- | --- | --- | --- |
| PPW 1 | $\Delta = -0.016$<br>[-0.101, 0.083] | $\Delta = -0.022$<br>[-0.109, 0.058] | $\Delta = 0.022$<br>[-0.084, 0.110] | **$\Delta = 0.124$**<br>**[0.006, 0.245]** | $\Delta = 0.064$<br>[-0.063, 0.197] | $\Delta = 0.109$<br>[0.063, 0.304] |
| PPW 2 | | $\Delta = -0.005$<br>[-0.068, 0.046] | $\Delta = 0.038$<br>[-0.061, 0.122] | **$\Delta = 0.140$**<br>**[0.005, 0.221]** | $\Delta = 0.080$<br>[-0.020, 0.197] | $\Delta = 0.125$<br>[-0.003, 0.277] |
| PPW 3 | | | $\Delta = 0.044$ | **$\Delta = 0.146$**<br>**[0.075, 0.221]** | **$\Delta = 0.085$**<br>**[0.010, 0.176]** | **$\Delta = 0.131$**<br>**[0.017, 0.270]** |
|  |  |  | [-0.009,<br>0.087] |  |  |  |
| PPW 4 | | | | **$\Delta = 0.102$**<br>**[0.019, 0.184]** | $\Delta = 0.042$<br>[-0.030,<br>0.137] | $\Delta = 0.087$<br>[-0.020,<br>0.228] |
| PPW 5 | | | | | **$\Delta = -0.060$**<br>**[-0.116, -**<br>**0.004]** | $\Delta = -0.015$<br>[-0.114,<br>0.104] |
| PPW 6 | | | | | | $\Delta = 0.045$<br>[-0.019,<br>0.120] |
| $\Delta = ICC_{row} - ICC_{column}$, presenting the ICC difference. A positive $\Delta$ indicates higher reliability for the row measurement, and vice versa for a negative $\Delta$. A 95% CI including 0 indicates no significant difference; conversely, a CI excluding 0 indicates a significant difference (highlighted cells). | | | | | | |

**Table 8.** Pairwise ICC differences among the seven PPW thickness measurement approaches normalized by C2C4 length.

| Approach | PPW 2 | PPW 3 | PPW 4 | PPW 5 | PPW 6 | PPW 7 |
| --- | --- | --- | --- | --- | --- | --- |
| PPW 1 | $\Delta = 0.019$<br>[-0.065,<br>0.109] | $\Delta = -0.013$<br>[-0.097,<br>0.059] | $\Delta = 0.016$<br>[-0.074,<br>0.090] | **$\Delta = 0.149$**<br>**[0.039, 0.261]** | **$\Delta = 0.106$**<br>**[0.003, 0.222]** | **$\Delta = 0.190$**<br>**[0.060, 0.350]** |
| PPW 2 | | $\Delta = -0.031$<br>[-0.086,<br>0.019] | $\Delta = 0.002$<br>[-0.092,<br>0.082] | **$\Delta = 0.131$**<br>**[0.057, 0.221]** | $\Delta = 0.087$<br>[-0.004,<br>0.213] | **$\Delta = 0.172$**<br>**[0.038, 0.356]** |
| PPW 3 | | | $\Delta = 0.029$<br>[-0.018,<br>0.075] | **$\Delta = 0.162$**<br>**[0.091, 0.237]** | **$\Delta = 0.119$**<br>**[0.046, 0.211]** | **$\Delta = 0.203$**<br>**[0.073, 0.358]** |
| PPW 4 |  |  |  | **$\Delta = 0.133$**<br>**[0.043, 0.222]** | **$\Delta = 0.090$**<br>**[0.012, 0.192]** | **$\Delta = 0.174$**<br>**[0.057, 0.314]** |
| PPW 5 | | | | | $\Delta = -0.044$<br>[-0.104, -<br>0.023] | $\Delta = 0.041$<br>[-0.075,<br>0.174] |
| PPW 6 |  |  |  |  |  | **$\Delta = 0.084$**<br>**[0.013, 0.165]** |
| $\Delta = ICC_{row} - ICC_{column}$, presenting the ICC difference. A positive $\Delta$ indicates higher reliability for the row measurement, and vice versa for a negative $\Delta$. A 95% CI including 0 indicates no significant difference; conversely, a CI excluding 0 indicates a significant difference (highlighted cells). | | | | | | |

Pre- and post-swallow comparisons were performed within the same selected bolus trial for each MBS examination. For each rater, the seven PPW thickness and cervical vertebral reference lengths measurements obtained from the pre-swallow resting frame were paired with the corresponding seven measurements (without normalization) from the post-swallow resting frame of the same video, and paired t-tests were performed separately for each PPW measurement approach. These paired comparisons were first conducted separately for each rater. A third paired comparison was then performed using the mean values of the two raters’ measurements at each time point, with the mean pre-swallow value paired with the mean post-swallow value from the same video for each PPW measurement approach. The paired t-tests tested whether the mean difference in PPW thickness between the pre- and post-swallow resting frames differed from zero. The p-values of all the comparisons are listed in the Table 9. No significant differences in PPW thickness were identified between the pre-swallow and post-swallow resting frames for any of the seven measurement approaches. This finding was consistent for measurements from Rater 1, Rater 2, and the mean values of the two raters. PPW 4 in Rater 1 had the lowest p-value (p = 0.0600), approaching but not reaching statistical significance at the level of 0.05.

**Table 9.** Paired comparisons of pre-swallow and post-swallow PPW thickness measurement.

| Options | *p*-value of Paired *t*-test<br>(PPW thickness measurement pre- vs. post- swallowing) |  |  |
| --- | --- | --- | --- |
|  | PPW thickness Measured<br>by Rater 1 | PPW thickness Measured<br>by Rater 2 | Mean PPW thickness<br>(Rater 1 + Rater 1)/2 |
| PPW 1 | 0.2188 | 0.8792 | 0.5300 |
| PPW 2 | 0.2667 | 0.2989 | 0.9679 |
| PPW 3 | 0.9808 | 0.3618 | 0.4788 |
| PPW 4 | 0.0600 | 0.9509 | 0.3146 |
| PPW 5 | 0.2216 | 0.5657 | 0.2385 |
| PPW 6 | 0.8410 | 0.6063 | 0.7652 |
| PPW 7 | 0.5724 | 0.1744 | 0.3777 |
| C2-C4 length | 0.3661 | 0.2855 | 0.9248 |
| C3 length | 0.3455 | 0.4163 | 0.9047 |

## Discussion

This study first conducted a scoping review to characterize existing approaches and methodological attributes of PPW thickness measurement on videofluoroscopic imaging. These prior studies suggest that PPW assessment is supported by an established methodological foundation, with broad convergence on anatomically anchored linear measurements, indicating that the construct itself is measurable and reproducible in principle. The more important methodological gap lies in how this construct is operationalized across studies. Informed by these studies, we developed a computer-assisted framework for PPW thickness measurement and evaluated the reliability of the independent raters across a range of conditions. Our findings support the feasibility of quantifying PPW thickness as a structural measure for treatment-related tissue changes. Particularly for patients with HNC, this measure may facilitate characterization of the development and severity of treatment-induced internal lymphedema following radiation therapy.

Bolus consistency and volume represent methodological variables that warrant consideration when selecting a bolus condition for PPW thickness measurement from multiple bolus-trials of a MBS, particularly with respect to whether the selected condition provides a representative assessment of PPW morphology. However, there is currently no convincing peer-reviewed evidence demonstrating that independent manipulation of either bolus consistency or bolus volume alters PPW thickness on videofluoroscopic imaging. Most studies adopted bolus conditions from established MBS protocols without providing an explicit rationale specific to PPW thickness measurement. Mancopes et al. compared resting PPW thickness between a cup sip of thin liquid and a teaspoon of mildly thick liquid and expected to maximize the likelihood of detecting consistency-related differences in PPW measures, however, no significant measurement differences was found between the two bolus conditions(36). In this study, a 5-mL thin liquid bolus was selected with reference to the bolus condition reported by Turcotte MC et al.(31) and its incorporation into the standardized MBS protocol at MD Anderson Cancer Center. We believe this bolus condition unlikely to result in substantial differences in PPW thickness measurement compared with bolus conditions adopted in other studies, such as a 20-ml thin bolus in the study of Leonard & Belafsky (33)

For the assessment of internal lymphedema, PPW thickness does not require frame-by-frame measurement across the entire swallow, as the primary objective is to quantify structural tissue thickness rather than dynamic PPW movement. Therefore, selecting a relatively static frame is critical for reliable assessment. Accordingly, this study employed a frame-selection approach consistent with previous studies, using a frame representing swallow rest, and for the pre-swallow condition, the resting state was equivalent to the bolus-hold position (15, 21, 31, 34). More specifically, the rest point in VFSS was defined as the point at which the pyriform sinuses reached their lowest position. However, no direct comparison has evaluated pre-swallow and post-swallow PPW thickness within the same swallowing trial to determine whether the two resting conditions yield different measurement values. In this study, we compared the measurements in these two time points within the same trial, and the analysis did not find statistical evidence of a difference between them, suggesting that, when PPW thickness is assessed under a relatively static resting condition, whether the selected frame before or after the swallow may not be a major source of measurement variability.

A distinct difference in interrater reliability was observed according to the vertebral level used for PPW measurement. Without normalization, the C2- and C3-based approaches (PPW 1-4) demonstrated ICCs ranging from 0.70 to 0.78, whereas reliability decreased for the C4-based approaches (PPW 5-7), with ICCs ranging from 0.51 to 0.62, as shown in Table 4. This cervical level-dependent difference persisted after normalization by either the C2-C4 or C3 reference length, as shown in Table 6. This pattern may reflect reduced delineation of the PPW boundary at the C4 level, where lower image contrast can make the wall margin more challenging to identify consistently between raters. In some cases, even at resting status, the lowest position of the pyriform sinus did not extend below the C4 level(37). Under these conditions, the PPW boundary around C4 became blurred on fluoroscopic imaging, as shown by the red arrow in Figure 4(a), in contrast, the PPW boundary at the C2 and C3 levels was well visualized on this frame. For the raters, a practical approach was to identify frames in which the bolus passes the C4 level and approached or entered the upper esophageal sphincter, as these frames often provide clearer delineation of the PPW boundary at C4, as shown by the red arrow in Figure 4(b). The clearly visualized PPW contour can then be used as a reference to approximate the corresponding boundary location on the resting frames. However, this estimation process may introduce additional interrater variability.

**Figure 4.**
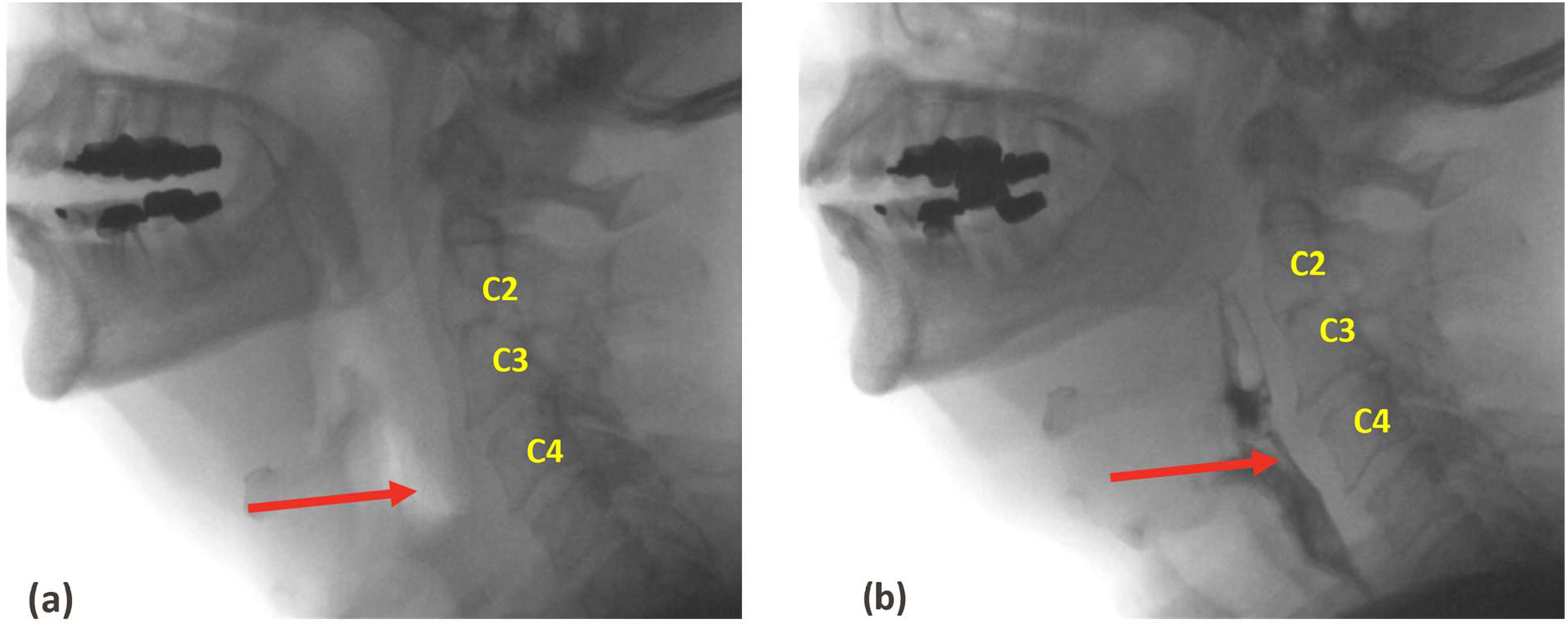
Example of limited PPW visualization at the C4 level. (a) Resting frame with limited PPW delineation near C4; (b) Bolus-passage frame showing clearer delineation of the PPW near C4.

A slightly lower ICC was also observed for PPW 1, which was based on the anterior-inferior corner of C2 (ICC = 0.70 without normalization). This may be related to the hook-like inferior contour of C2 that was present in some cases, also presented in Figure 4, which can make precise identification of the anterior-inferior corner more challenging for raters. However, the ICC value observed for PPW 1 suggests that this anatomical variation did not substantially compromise measurement reliability. The C3-based approaches (PPW 2-4) provided the most reliable measurements, likely reflecting the consistently favorable visualization of the PPW boundary at this level. Among these approaches, PPW 3, which was based on the midpoint of C3, demonstrated the highest interrater reliability both before and after normalization. Notably, PPW 3 requires accurate calculation of the C3 midpoint, which may not be practical in clinical settings or research workflows where rapid manual measurement is required. Alternatively, PPW 4, based on the anterior-inferior corner of C3, may represent a practical measurement, as adopted by Turcotte *et al* (31). Its interrater reliability was only slightly lower than that of PPW 3 before normalization (ICC = 0.76 vs. 0.78), while allowing the measurement to be anchored directly to an identifiable anatomical landmark.

As introduced above, a fundamental limitation of MBS imaging is that patient-to-detector distance and imaging geometry cannot be held constantly across examinations, preventing direct interpretation of image-based distances in physical units such as mm or cm. Consequently, pixel-based measurements are not directly comparable across examinations or longitudinal time points, and a reference scale is required for standardized PPW thickness measurement. In our scoping review, most studies (5/9) used an external reference of known dimensions for calibration. Disc-shaped radiopaque markers, such as a coin, were most commonly used. This approach is not specific to PPW thickness and has also been applied to other anatomical measurements derived from VFSS imaging (38–40). External references, however, have several practical and methodological limitations. Their implementation may be difficult in routine clinical MBS examinations and more feasible in well-controlled research or clinical-trial settings. In addition, an external marker does not necessarily lie in the same imaging plane as the PPW, which is a deeper anatomical structure. Because fluoroscopic magnification varies with object-to-detector distance, pixel-to-physical-unit conversion using an external marker may be affected by depth-dependent magnification (41, 42). In lymphedema assessment, longitudinal change in internal tissue thickness may be more informative than absolute thickness at a single examination. An internal anatomical reference may therefore be better suited for standardized relative measurement across examinations, time points, and individuals(43). Supporting this approach, Jones-Rastelli et al. found no within-subject change between timepoints in penny diameter expressed relative to C2-C4 distance(34).

Among the two internal anatomical references evaluated, the C3 length demonstrated lower interrater reliability than the C2-C4 length (ICC = 0.72 vs. 0.88). This difference was further reflected in the normalized PPW measurements, for which C3-based normalization consistently yielded lower interrater reliability than C2-C4-based normalization, as shown in Table 5. One possible explanation is that longer landmark-determined distances are less sensitive to a similar absolute magnitude of landmark-localization error when the imaging landmark is well-defined (distance-error variance is a constant), resulting in smaller relative measurement error and potentially greater reliability(44, 45).

In this study, the computer-assisted and semi-automated framework was designed to establish the quantified PPW measurement and reduce the extent to which rater-dependent decisions propagate through the measurement process. Rater input was retained for steps requiring anatomical interpretation, particularly identification of cervical vertebrae corners landmarks and the PPW boundary, while other components of the measurement are objectively calculated using predefined geometric computations. In this way, the framework limits the rater’s discretion to the components that require visualization judgment while standardizing the remaining steps of the measurement process. This collaboration between human interpretation and computational constraints was intended to reduce subjective variability and improve measurement consistency and reproducibility. Also, this framework can be readily integrated with other image-analysis methods, potentially paving the way for more fully automated approaches to PPW measurement.

## Limitation

This study focused primarily on measurement reliability and did not directly establish the validity of PPW thickness as a marker of internal lymphedema. Although videofluoroscopy in MBS is widely regarded as a gold standard for assessment of swallowing function, it is not necessarily a standard for measurement of PPW thickness. Some prior studies have provided indirect validity evidence for fluoroscopic PPW measures. Evangelista et al. reported associations between increased PPW thickness and pyriform sinus retention, higher PAS scores, and post-deglutitive aspiration, while Jones-Rastelli et al. identified a relationship between PPW thickness and pharyngeal residue(15, 34). Turcotte et al. demonstrated longitudinal increases in PPW thickness following chemoradiation(31), and Gregor et al. provided evidence of responsiveness to an intervention targeting head and neck lymphedema(30). Further validation of PPW thickness measurements may require comparison with an independent imaging modality capable of more directly characterizing tissue thickness. Endoscopic assessment, although commonly used for evaluation of internal lymphedema, primarily visualizes the mucosal surface and does not permit direct measurement through the PPW tissue. CT (Computed Tomography) or MRI (Magnetic Resonance Imaging) may therefore provide more appropriate complementary reference measurements for future validation studies.

Another limitation relates to the definition of measurement direction. The measurement directions used in this study were defined perpendicular to the local anterior surface of the corresponding vertebral body, following the approach described by Watanabe *et al*(32): PPW 2-4 were referenced to C3 surface and PPW 5-7 to C4 surface. This approach assumes that the adjacent PPW boundary is approximately parallel to the local vertebral surface and also allows the measurement direction to reflect angular changes in cervical curvature across vertebral levels. However, when cervical curvature is limited, the cervical spine can alternatively be approximated by a single longitudinal axis, such as the line defined by the C2-C4 anterior-inferior landmarks, as used by Mancopes *et al* (36). In that case, PPW thickness could be measured perpendicular to this common C2-C4 axis rather than to the local C3 or C4 surface. This alternative orientation was not evaluated in the present study. We speculate that, when the angular difference between the local vertebral surface and the C2-C4 axis is minimal, the resulting PPW measurements and their reliability may be similar between the two approaches.

## Conclusion

This study provides a methodological foundation for quantitative measurement of PPW thickness on MBS. By integrating methodological evidence from scoping review of previous studies with an empirical evaluation of measurement reliability, the proposed framework formalizes PPW thickness as a reproducible structural measure while reducing variability introduced by subjective measurement decisions. This approach extends the quantitative use of MBS beyond swallowing function alone and provides a basis for incorporating structural tissue assessment into longitudinal evaluation of treatment-related structural changes in patients with HNC.

## Data Availability

All data produced in the present study are available upon reasonable request to the authors

